# Multimodal Non-Invasive Biomarker Characterization of Structural and Functional Alterations in ADSS1 Myopathy

**DOI:** 10.64898/2026.01.25.26344324

**Authors:** Merve Koç Yekedüz, Raquel van Gool, Hanne van der Heijden, Buket Sonbas Cobb, Nehal Shah, Georgina Johnson, Cara A Timpani, Julie Shulman, Vanessa Rameh, Evan E Hsu, Courney LeSon, Pui Y Lee, Adam P. Vogel, Walla Al-Hertani, Hyung Jun Park, Emma Rybalka, Seward B. Rutkove, Jaymin Upadhyay

**Affiliations:** Department of Anesthesiology, Critical Care and Pain Medicine, Boston Children’s Hospital, Harvard Medical School, Boston, MA, USA; Department of Pediatric Metabolism, Ankara University, Faculty of Medicine, Ankara, Türkiye; Rare Diseases Application and Research Center, Ankara University, Ankara, Türkiye; Department of Neurology, Mental Health and Neuroscience Research Institute, Maastricht University, Maastricht, Limburg, The Netherlands; Department of Neurology, Beth Israel Deaconess Medical Center, Harvard Medical School, Boston, Massachusetts, USA; Department of Electrical and Electronic Engineering, Harran University, Sanliurfa, Türkiye; Department of Radiology, Brigham and Women’s Hospital, Harvard Medical School, Boston, MA, USA; Department of Audiology and Speech Pathology, The University of Melbourne, Parkville, Victoria, Australia; Inherited and Acquired Myopathies Program, Australian Institute for Musculoskeletal Science, St Albans, Victoria, Australia; Department of Medicine-Western Health, Melbourne Medical School, The University of Melbourne, St Albans, Victoria, Australia; Department of Physical and Occupational Therapy, Boston Children’s Hospital, Boston, MA, USA; Department of Radiology, Boston Children’s Hospital, Harvard Medical School, Boston, MA, USA; Division of Immunology, Boston Children’s Hospital, Harvard Medical School, Boston, MA, USA; Redenlab Ltd, Melbourne, Victoria, Australia; Division of Metabolic Disorders, Children’s Hospital of Orange County (CHOC®), Rady Children’s Health, Orange, CA, USA; Department of Neurology, Yonsei University College of Medicine, Gangnam Severance Hospital, Seoul, Republic of Korea

**Keywords:** Adenylosuccinate synthetase (ADSS1) myopathy, muscle, biomarkers, proteomics, speech, magnetic resonance imaging

## Abstract

Adenylosuccinate synthetase 1 (ADSS1) myopathy is an ultra-rare disease characterized by progressive muscle dysfunction. The objective of this investigation was to employ a non-invasive biomarker approach to phenotype (fine-)motor skills, speech production and cognition in adults with ADSS1 myopathy. Five individuals with ADSS1 myopathy and five age-sex-matched healthy controls (HCs) underwent a comprehensive multimodal evaluation. Assessments included, (i) evaluation of motor performance, (ii) speech production and cognitive test batteries, (iii) patient-reported outcomes, (iv) electrical impedance myography (EIM), (v) musculoskeletal magnetic resonance imaging (MRI) and (vi) plasma proteomics. ADSS1 participants vs. HCs demonstrated reduced performance on the 9-Hole Peg and grip strength tests as well as lower self-reported mobility. Speech production analysis revealed asthenia (p=0.02), lower intelligibility (p=0.008), and worse voice quality during the sustained vowel task (p=0.03) in the ADSS1 cohort. Cognitive functioning remained unaffected in patients with ADSS1. On EIM, ADSS1 participants vs. HCs, demonstrated a pattern of higher resistance and lower reactance and phase across upper- and lower-extremity measurements, indicative of poorer muscle health, with large effect sizes (Cliff’s δ=0.5-0.9). MRI revealed intramuscular fat infiltration, particularly in posterior compartments of the upper leg (e.g., biceps femoris). Proteomics indicated reduced (p=0.04) Neurotrophin-3 (NTF3; implicated in neuronal development, survival and differentiation) levels in the ADSS1 cohort relative to HCs. Lower NTF3 levels associated with poorer performance on hand-motor tasks as well as higher resistance and lower reactance and phase on EIM. This study highlighted the value of multimodal phenotyping for quantifying disease expression and advancing monitoring strategies in ADSS1 myopathy.

**Take-home message:** This multimodal investigation demonstrates that integrating electrical impedance myography with quantitative motor, speech, musculoskeletal imaging, and proteomic assessments provides a sensitive and non-invasive research framework for capturing neuromuscular dysfunction and functional disease burden in patients with ADSS1 myopathy, thereby supporting the current biomarker strategy for refined phenotyping and longitudinal disease monitoring in this ultra-rare condition.

## INTRODUCTION

Adenylosuccinate synthetase 1 (ADSS1) myopathy (formerly called ADSS1-like 1 (ADSSL1) myopathy) is an ultra-rare, hereditary neuromuscular disorder caused by biallelic pathogenic variants in *ADSS1*, an enzyme that plays a critical role in the purine nucleotide cycle (PNC) and AMP biosynthesis within skeletal muscle.^1-7^ Within the United States there are 5 known cases of ADSS1 myopathy to date, with larger cohorts in India, South Korea, and Japan.^2,4,8,9^ There are no curative treatments for ADSS1 myopathy^7^. Disruption of ADSS1-dependent metabolic processes is believed to impair myocyte viability, compromise myoblast proliferation, and alter glycolytic and oxidative pathways, ultimately contributing to progressive muscle weakness and functional decline.^8,10^ Clinically, ADSS1 myopathy exhibits considerable heterogeneity.^2,4,8,9^ Patients may present with distal or proximal limb weakness, exercise intolerance, fatigability, dysphagia, gait impairment, or a combination of musculoskeletal and functional difficulties. Patients with this ADSS1 myopathy also frequently exhibit dysarthria and fatty replacement of the tongue.^2,5^ Although the number of reported cases has increased in recent years, understanding of the disease continues to evolve, while natural history data remain sparse.^11-14^

Presently, diagnostic and monitoring approaches in ADSS1 myopathy rely on genetic testing, standard neuromuscular functional assessment batteries, musculoskeletal magnetic resonance imaging (MRI), electromyography, serum biomarkers, physical examination, and, in select cases, muscle biopsy. While these are valuable methods, they have limitations. For example, some non-ambulatory patients may not be able to perform certain motor tasks, while young children with ADSS1 myopathy may require sedation to undergo MRI. Patients may also experience significant difficulty when lying flat during imaging procedures (i.e., individuals dependent on respiratory assistive devices). Additionally, some modalities are resource-intensive, invasive, or may be limited in their ability to detect early or subclinical changes in muscle health or function. An unfortunate outcome of current methodologies for clinical course assessment, considering applicability, accessibility, or sensitivity, is inadequate monitoring of disease mechanisms or core symptoms in ADSS1 myopathy. This can lead to delays in diagnosis or subsequent implementation of any form of clinical interventions (e.g., physical therapy or occupational therapy). Given these unmet needs, the aim of this investigation was to identify objective, reliable, flexible, and sensitive clinical methods that can be applied to a broad range of adult patients with ADSS1 myopathy for purposes of detecting compromised muscle integrity and monitoring core symptoms, mainly, deficits in (fine-)motor performance and dysarthria.

In prior work, electrical impedance myography (EIM) was utilized to identify muscle abnormalities in patients with ADSS1 myopathy^11^, while a more recent multinational effort focused on further comprehending the personal challenges faced by patients with ADSS1 myopathy and their caregivers^9^. Guided by these earlier investigations, the objective of this prospective study was to undertake more extensive phenotypic characterization of ADSS1 myopathy and explore the utility of non-invasive biomarkers in capturing both structural and functional alterations associated with the disease. Moreover, although clinical features of ADSS1 myopathy have focused predominantly on motor impairment or skeletal muscle abnormalities, the broader functional phenotype including, for example, dysarthria, remains largely unexplored. Thus, a multimodal approach integrating muscle imaging, electrophysiological assessment, neurofunctional characterization, and plasma proteomics alongside an exploratory muscle transcriptomics analysis^3^, was used to advance our understanding of this condition and identifying measurable indicators of disease burden in ADSS1 myopathy.

## METHODS

### Study Participants

This study was approved by the Boston Children’s Hospital (BCH) Institutional Review Board (IRB-P00040784) and adhered to Helsinki criteria for human subject research. This study prospectively enrolled individuals with genetically confirmed ADSS1 myopathy (identified through the ADSS1 myopathy disease registry) and age- and sex-matched healthy controls (HCs). Controls had no neuromuscular, systemic disease or acute injury or illness. Demographic and anthropometric characteristics were collected at enrollment. All participants provided written informed consent prior to study participation.

### Motor and Functional Assessments

Upper-extremity motor performance was evaluated using grip strength and the 9-Hole Peg tests. While the grip strength test was implemented as a proxy measure of overall muscle strength and physical health, the 9-Hole Peg tests enabled an evaluation of fine-motor skills and coordination^15,16^. One patient with ADSS1 could not complete 9-Hole Peg tests due to disease severity. Functional status and patient-reported outcomes were assessed using Patient-Reported Outcomes Measurement Information System (PROMIS-short forms version 7a) measures for Mobility, Fatigue, Pain, Cognitive Function, Anxiety, Depression, and Psychological Stress^17^. Quality of Life in Neurological Disorders (Neuro-QoL) domains—Communication, Sleep Disturbance, and Swallowing Difficulty—were also administered^18^. Raw scores were converted to T-scores for all PROMIS and Neuro-QoL questionnaires.

### Speech Production

ADSS1 expression is prominent in oropharyngeal musculature which may explain symptomatic dysarthria.^5^ To assess this aspect of the disease, a battery probing motor and cognitive aspects of speech was utilized.^19^ Participants completed a speech battery that included (i) sustained vowel /a/ production for 5 seconds, (ii) rapid and precise syllable repetition (/pa-ta /) (diadochokinesis, DDK) for 10 seconds, (iii) reading a passage (137 syllables), (iv) a picture description task, and (v) monologue for 1-2 minutes. The sustained vowel, syllable repetition, and reading tasks were repeated twice to minimize the impact of task unfamiliarity and increase measurement reliability.^20,21^ Speech was recorded using Redenlab® Online software. Speech samples were analyzed objectively using acoustic analysis and natural language processing and subjectively via expert perceptual judgement by a blinded speech-language pathologist (GJ). Voice quality was measured via cepstral peak prominence and harmonics to noise ratio, vocal control via variability of sound pressure level and fundamental frequency, and timing via syllable duration, articulation rate and voice onset time.^22-25^ Speech and voice features were rated using the GRBAS scale,^26^ direct magnitude estimation (DME) for intelligibility and naturalness,^27^ speech subsystem functioning (e.g., articulation, phonation, resonance), and diadochokinetic rates.^28^ The GRBAS scale assesses the presence and severity of vocal disorders across five domains: Grade (G) or overall vocal disorder, Roughness (R), Breathiness (B), Asthenia (A) or weakness, and Strain (S) using a scale from 0 to 4 (0 = subclinical/no issue, 1 = mild, 2 = moderate, 3 = severe, 4 = profound) based on sustained phonation and monologue or picture description tasks. DME uses an anchor score of 100 representing mild dysarthria. Scores are allocated on a continuous scale on measures of intelligibility and naturalness by comparing the performance of speakers to that of the anchor. Scores lower than 100 represent less intelligible and less natural sounding speech, with 100 as the maximum score. Diadochokinetic performance was evaluated on the syllable repetition task for speed, consistency, and accuracy on a scale of 0-4.

### Cognition

In a prior multinational investigation of ADSS1 patients and caregivers, cognitive difficulties were noted by a subset of individuals.^9^ Therefore, cognitive performance was assessed using the NIH Toolbox Cognitive Battery. Executive function was evaluated through the Dimensional Change Card Sort (DCCS), Flanker Inhibitory Control task, and List Sorting Working Memory task. Language-related skills were assessed via Oral Reading and Picture Vocabulary measures. General cognition was quantified using Crystallized, Fluid, and Total Composite scores. Standardized scoring procedures provided age-adjusted performance values for all completed tasks.

### Electrical Impedance Myography

EIM measurements were acquired in accordance with methodologies parallel to those previously described.^11,29^ Briefly, a portable EIM device (mScan, Myolex Inc., Brookline, MA, USA) along with an accompanying iPad (Apple Inc., Cupertino, CA, USA) with EIM data acquisition software was utilized. EIM was performed on bilateral upper and lower extremity muscles, while participants remained in a relaxed, supine position. Upper extremity muscles assessed included: middle deltoid, biceps brachii, wrist extensors, wrist flexors. Lower extremity muscle included: rectus femoris and medial gastrocnemius. Measurements on each muscle group were repeated two to three times and traces were reviewed in real-time for artifacts, poor contact, or abnormal signal distortions.^30^ Data were collected at multiple frequencies (EIM scan frequency range: 1–10,000 kHz). Resistance, reactance, and phase were extracted at 100 kHz based on previous studies, as the capacitive nature of the muscle is greatest around this frequency.^30,31^ Each unilateral muscle group and each EIM trace underwent further quality assurance/quality control to identify any signal artifacts potentially missed during real-time data assessment. For each muscle, the average EIM trace of two to three measurements was generated and utilized for further EIM analysis.

### MRI

MRI data were acquired on a Siemens 3T Scanner (Siemens, Erlangen, Germany) at BCH in combination with a spine coil and body array flex coil and extender (4 element design) for non-contrast musculoskeletal MRI of the lower limbs, from the pelvic crest to the patella. A second musculoskeletal MRI acquisition was performed on the subject’s right lower leg using a 15 channel-knee coil. Dixon MRI was used to quantify fat fraction in key muscle groups by NS. Dixon MRI pulse sequence parameters are provided in **Supplemental Material.** Fat fractions were quantified at three levels in the thigh muscles, ranging from proximal to distal, with the most distal level corresponding with the level of EIM electrode array placement. For each level, anterior and posterior muscle compartments were manually segmented, and fat fraction values were quantified. Fat fraction quantification focused on the anterior (vastus lateralis, vastus intermedius, rectus femoris, vastus medialis) and posterior (biceps femoris, semitendinosus, semimembranosus) compartments. For whole-head non-contrast craniofacial MRI, a Siemens 3T Scanner (Siemens, Erlangen, Germany) with a 64-channel head coil was utilized to assess the masseter muscles by VR. T1- and T2-weighted pulse sequence parameters are provided in **Supplemental Material.** MRI acquisition was only feasible in three out of five patients with ADSS1 myopathy.

### Plasma Proteomics

Fluid biomarkers are clinically useful and inexpensive tools that potentially inform prognosis. Each study participant provided a blood sample (8-10 mL). Plasma was aliquoted, and frozen at −80°C. The Olink platform utilizes a well-characterized proximity extension assay (PEA), which allows for high-throughput, multiplex protein quantification with minimal sample volume (i.e., 1 µL of plasma was used per assay). Absolute concentrations (pg/mL) were determined using a calibrator system and pre-defined standard curves. All assays were performed using the Olink Q-100 benchtop instrument as described previously.^32^ Analytes in a customized Olink Flex panel targeted immune, neurological, cardiovascular, and musculoskeletal systems: GDNF, HIF1A, GDF2, IKBKG, CLEC4A, NTF3, ANGPT1, FGF21, KYNU, FGF23, DKK1, TLR1, TNFα, IL-6, IL-1β, IL-7, IL-17A, TNFSF10, CXCL8, IL33, and IFNG. To complement plasma proteomics findings, existing muscle transcriptomics data derived from Korean patients with ADSS1 myopathy (N=4) were explored to potentially identify common molecular markers across the two biological compartments (i.e., plasma and skeletal muscle).^3^

### Statistical Analysis

For all clinical measures, group comparisons between patients with ADSS1 and HCs was performed using a Mann Whitney U-test. Effect sizes (Cliff’s δ) were computed to characterize the magnitude of group differences. Effect sizes were categorized as follows: δ (negligible): < 0.2; δ (small): 0.2–0.3; δ (medium): 0.3–0.5; δ (large): > 0.5. For existing muscle transcriptomics data derived from patients with ADSS1, Cliff’s δ effect sizes were also calculated^3^. Correlations between parameters were assessed using Spearman’s correlation analysis. A significance threshold of p < 0.05 was applied.

## RESULTS

### Participant Overview

A total of five individuals with ADSS1 myopathy and five age- and sex-matched HCs were included in the study (**Table 1**). All patients with ADSS1 myopathy carried biallelic pathogenic variants in *ADSS1* (c.910G > A (p.Asp304Asn)), verified through prior clinical genetic testing. Two patients with ADSS1 myopathy were not ambulatory. Some participants had additional endocrine comorbidities. The distribution of sex was identical across groups (40% female, 60% male). Age, height, and weight did not significantly differ between the ADSS1 and control groups, indicating that the cohorts were demographically and anthropometrically comparable.

**Table 1.**
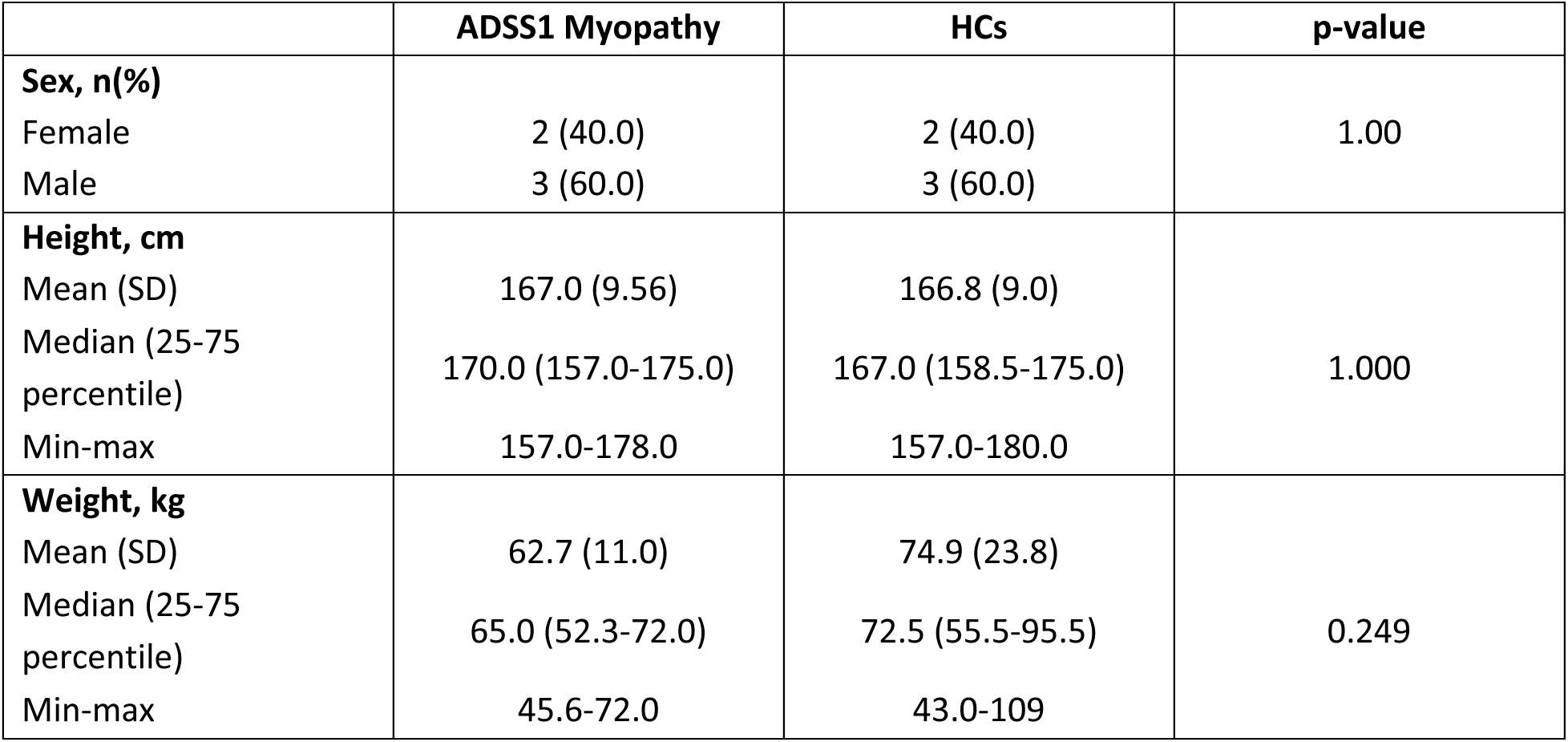
Demographic and Baseline Characteristics of Patients with ADSS1 Myopathy and HCs.

|  | **ADSS1 Myopathy** | **HCs** | **p-value** |
| --- | --- | --- | --- |
| **Sex, n(%)** |  |  |  |
| Female | 2 (40.0) | 2 (40.0) | 1.00 |
| Male | 3 (60.0) | 3 (60.0) |  |
| **Height, cm** |  |  |  |
| Mean (SD) | 167.0 (9.56) | 166.8 (9.0) | 1.000 |
| Median (25-75 percentile) | 170.0 (157.0-175.0) | 167.0 (158.5-175.0) |  |
| Min-max | 157.0-178.0 | 157.0-180.0 |  |
| **Weight, kg** |  |  |  |
| Mean (SD) | 62.7 (11.0) | 74.9 (23.8) | 0.249 |
| Median (25-75 percentile) | 65.0 (52.3-72.0) | 72.5 (55.5-95.5) |  |
| Min-max | 45.6-72.0 | 43.0-109 |  |

### Upper Extremity Motor Performance

Relative to HCs, participants with ADSS1 myopathy demonstrated poorer performance on the 9-Hole Peg Test with more impairment in the dominant vs. non-dominant hand **(Figure 1A).** Reduced bilateral grip strength was also observed in the ADSS1 myopathy cohort **(Figure 1B)**.

**Figure 1:**
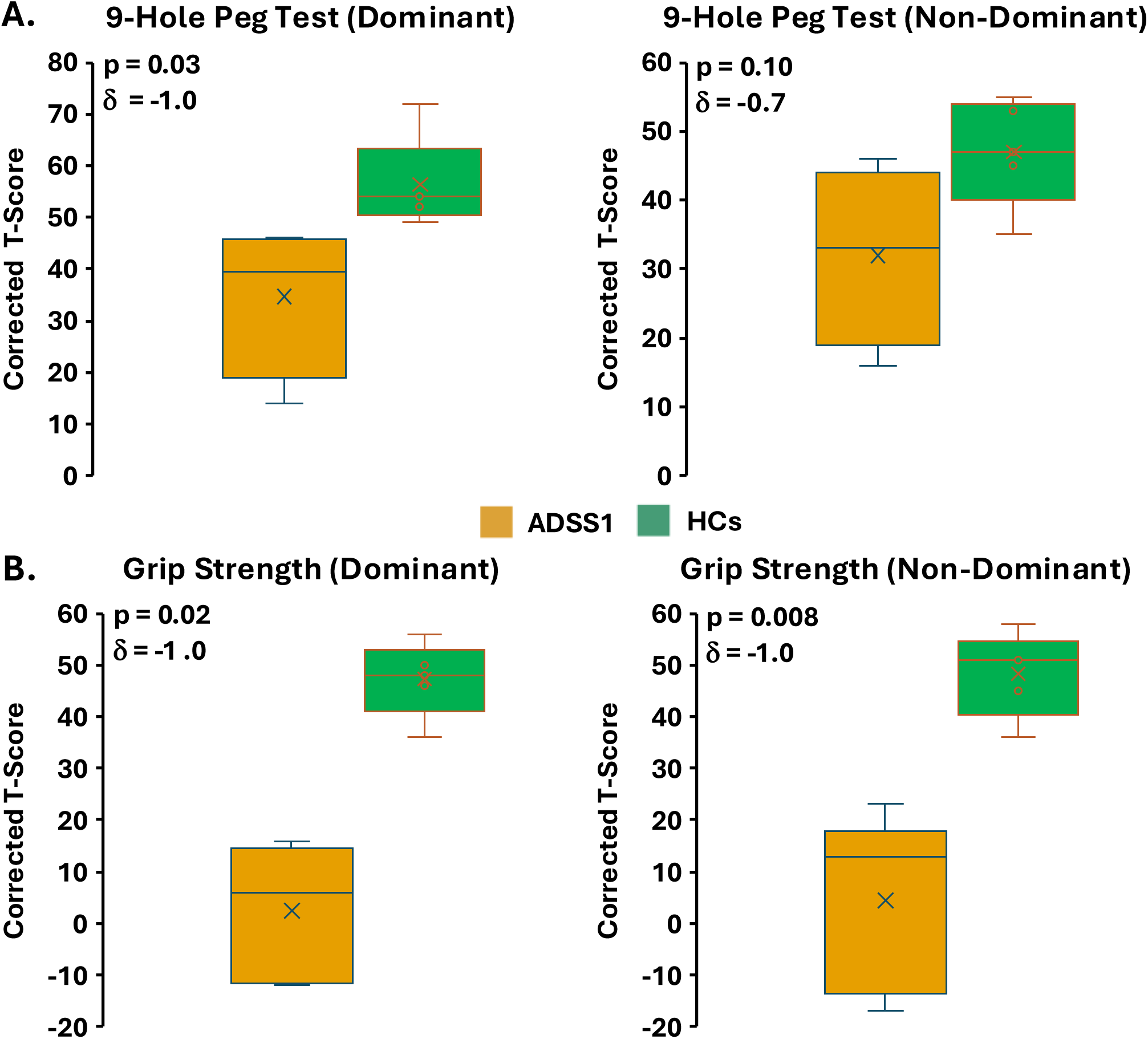
Reduced Upper Extremity Motor Performance. Patients with ADSS1 myopathy showed reduced performance one the 9-Hole peg test **(A)** and grip strength test **(B)** in the dominant and non-dominant hand.

### Patient-Reported Outcomes

In accord with upper extremity motor performance tests, mobility scores were significantly lower in the ADSS1 group (**Table 2**). Relative to HCs, patients with ADSS1 myopathy reported a trend of more fatigue and depressive symptoms. No notable group differences were observed across the remaining domains for PROMIS-based or Neuro-QoL-based measures.

**Table 2.** Patient-Reported Outcome Measures.

| | ADSS1 Myopathy | HCs | p-value | Cliff's $\delta$ |
| --- | --- | --- | --- | --- |
| **PROMIS** |  |  |  |  |
| Mobility | 27.8 ± 8.9 | 58.1 ± 4.7 | 0.008 | 1.0 |
| Cognitive Function | 53.2 ± 6.1 | 53.0 ± 11.1 | 1 | -0.04 |
| Psychological Stress Experience | 56.4 ± 5.1 | 48.6 ± 12.7 | 0.5 | 0.3 |
| Anxiety | 51.5 ± 11.4 | 47.3 ± 10.3 | 0.5 | 0.3 |
| Depressive Symptoms | 54.4 ± 9.4 | 44.8 ± 10.6 | 0.2 | 0.5 |
| Perceived Stress | 19.2 ± 4.6 | 19.6 ± 8.2 | 0.7 | 0.2 |
| **NeuroQol** |  |  |  |  |
| Fatigue | 50.1 ± 9.3 | 35.4 ± 8.4 | 0.06 | 0.8 |
| Swallowing Difficulties | 52.4 ± 16.5 | 41.0 ± 0 | 0.3 | 0.4 |
| Communication | 85.0 ± 28.3 | 100.0 ± 0 | 0.3 | -0.4 |
| Sleep Disturb | 45.8 ± 6.9 | 41.0 ± 9.7 | 0.4 | 0.3 |

### Speech Production and Cognitive Test Battery Assessment

Perceptual analysis of speech revealed vocal asthenia (weakness in the voice) and reduced intelligibility in patients with ADSS1 (**Figure 2A-B**). From acoustic analysis, the ADSS1 cohort presented with higher harmonics to noise ratio (HNR) during the sustained vowel task (/a/) (**Figure 2C**). A trend (p=0.056) of prolonged syllable duration for the syllable repetition task (‘pa-ta’) was also observed in patients with ADSS1 myopathy relative to HCs. From perceptual analysis of monologue and sustained vowel tasks, one older participant showed severe impairment in breathiness, overall voice quality, resonance, and intelligibility. The patient’s inability to produce consonant sounds may reflect excessive hypernasality. Two younger participants showed very mild or subclinical presentation of monoloudness, reduced stress, loudness decay, and mild voice quality issues. Acoustic analysis of sustained vowel and syllable repetition tasks revealed overall slower rate and impaired voice quality. The full set of perceptual and acoustic analysis results are provided in **Supplemental Table 1**. Lastly, radiological examination of whole-head, T1- and T2-weighted MRI collected in the three patients able to undergo MRI did not reveal abnormalities (**Figure 2D**). Specifically, signal abnormalities were not observed in either the left or right masseter muscle.

**Figure 2:**
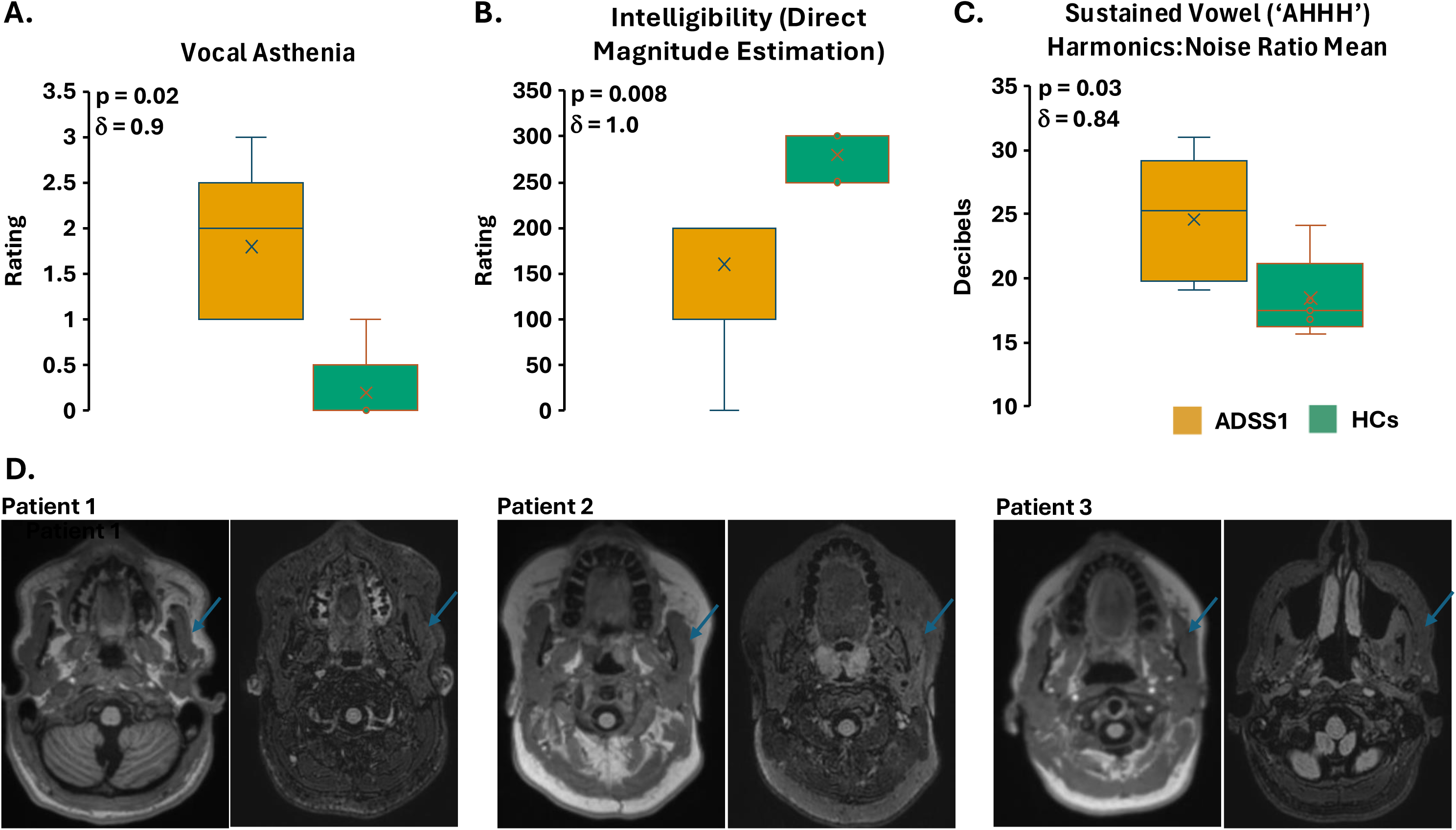
Characterization of Dysarthria in ADSS1 Myopathy. Perceptual and acoustic analyses were carried out on speech recordings. Weakness in voice **(A),** reduced intelligibility **(B)** and increased harmonics to noise ratio during the sustained vowel task **(C)** were observed in the ADSS1 cohort relative to HCs. Macroscopic signal abnormalities on MRI were not observed in the masseter muscle in the three patients with ADSS1 myopathy. See also **Supplemental Table 1**

Cognitive performance was intact in all five patients with ADSS1 myopathy. Significant differences between ADSS1 myopathy and HC cohorts or notable trends in were not present for executive function measures (i.e., DCCS, Flanker and List Sorting tasks), or for performance on language-related tasks (i.e., Oral Reading and Picture Vocabulary showed no group differences (p-value range: 0.2-0.8; Cohen’s *d* range: 0.1-0.5). All cognitive tasks were completed by all five patients with ADSS1 myopathy. The full set of cognitive performance results are provided in **Supplemental Table 2**.

### Evaluation of Muscle Health with EIM and Dixon MRI

ADSS1 participants showed consistent changes on EIM parameters, phase, resistance, and reactance at 100 kHz (**Figure 3**). Relative to HCs and for combined upper and lower extremity EIM assessments, patients with ADSS1 myopathy showed decreased phase and reactance values, while resistance was increased. This pattern, which is consistent with earlier reports and indicative of impaired muscle health^11^, was evident at the single-subject level, and also, in upper and lower extremities separately **(Supplemental Figure 1).** Dixon MRI was utilized to quantify muscle fat fraction in the lower limbs, specifically, the anterior and posterior muscle groups at 3 levels in the upper legs as well as in the right lower leg (**Figure 4**). Fat fraction in the upper leg for one patient with ADSS1 was not calculated due to technical issues during acquisition of Dixon MRI. All patients showed fat infiltration of upper and lower leg muscle tissue and specifically, higher fat fraction values in posterior versus anterior muscle groups. Multisequence MRI (i.e., T1-weighted, T2-weighted, and Short Tau Inversion Recovery MRI) of the upper leg (pelvic crest to patella for 3 patients with ADSS1 myopathy are shown in **Supplemental Figure 2**.

**Figure 3:**
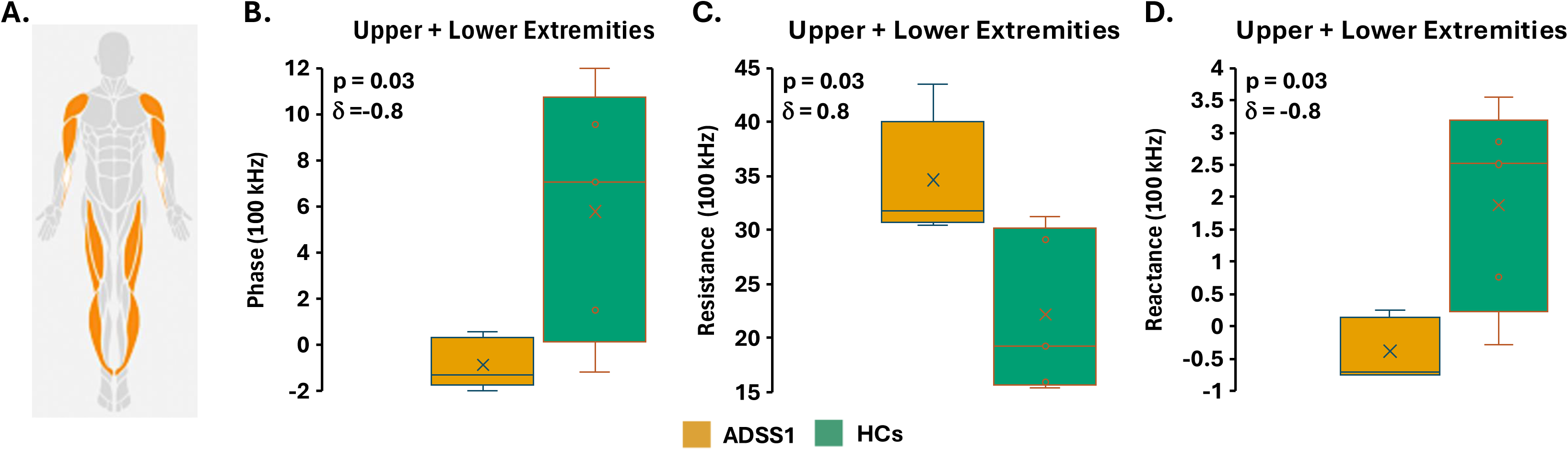
Compromised Muscle Health in ADSS1 Myopathy. EIM data was collected in multiple upper extremity and lower extremity muscle groups **(A)**. Muscle phase, resistance, and reactance at 100 kHz. Here, datapoints from upper and lower extremity muscle groups were combined. Lower phase **(B)**, higher resistance **(C)**, and lower reactance **(D)** values in the ADSS1 cohort were observed. See also **Supplemental Figure 1**.

**Figure 4:**
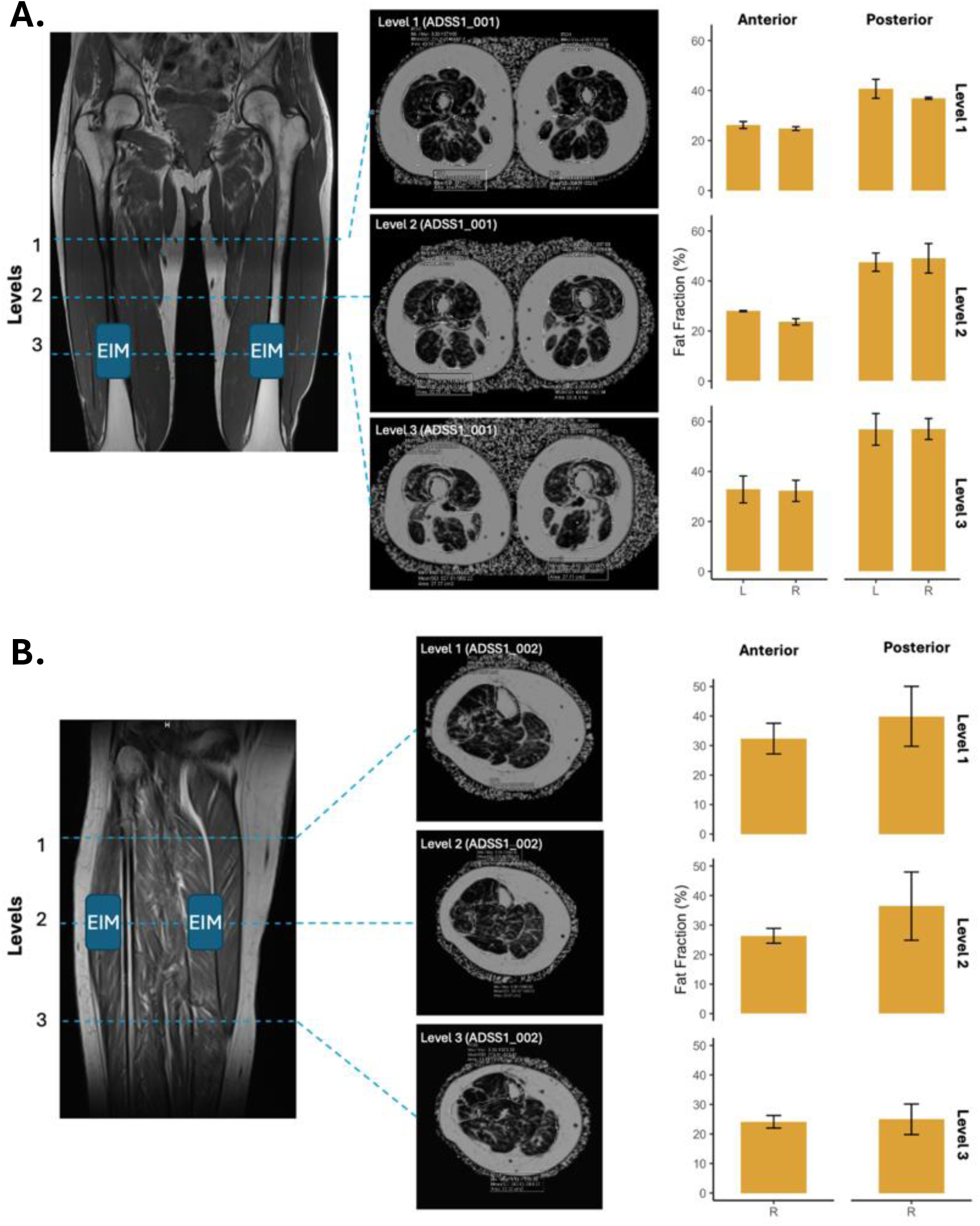
Quantification of Intramuscular Fat Infiltration. Three patients with ADSS1 myopathy were able to undergo MRI of the lower limbs. Dixon MRI was performed in the upper leg from the pelvic crest to patella **(A)**, and separately, in the right lower leg **(B)**. A pattern of higher levels of fat infiltration in posterior vs. anterior muscle groups in upper as well as lower legs was present. Additional multisequence MRI through the upper leg is provided in **Supplemental Figure 2**.

### Plasma Proteomics and Muscle Transcriptomics

Scoping for potential blood biomarkers of ADSS1 myopathy, we assayed a curated protein panel representing disease-relevant physiological systems involvement using Olink proteomics. Of the 21 proteins assayed (**Figure 5A**), only one was significantly changed (downregulated) – NTF3 (neurotrophin-3, p=0.04, δ=-0.8), which promotes axon regeneration, neuromuscular junction transmission and has anti-inflammatory actions in the CNS. However, based on moderate-large Cliff’s δ effect size (>0.3), 2 proteins were increased in ADSS1 patient plasma relative to HC (Interleukin-6 (IL-6); p=0.2, δ=0.5 and toll-like receptor-1 (TLR1); p=0.3; δ=0.4) and all were related to inflammation and immune response. IL-6, a pro-inflammatory cytokine and energy sensitive myokine, was most elevated (Cliff’s δ=0.5). Additionally, Inhibitor of Nuclear Factor Kappa B Kinase Regulatory Subunit Gamma (IKBKG) showed a trend of reduced (p=0.09, δ=-0.68) expression in the ADSS1 cohort. All other plasma proteomic markers showed negligible to medium effect sizes (**Supplemental Table 3**).

**Figure 5:**
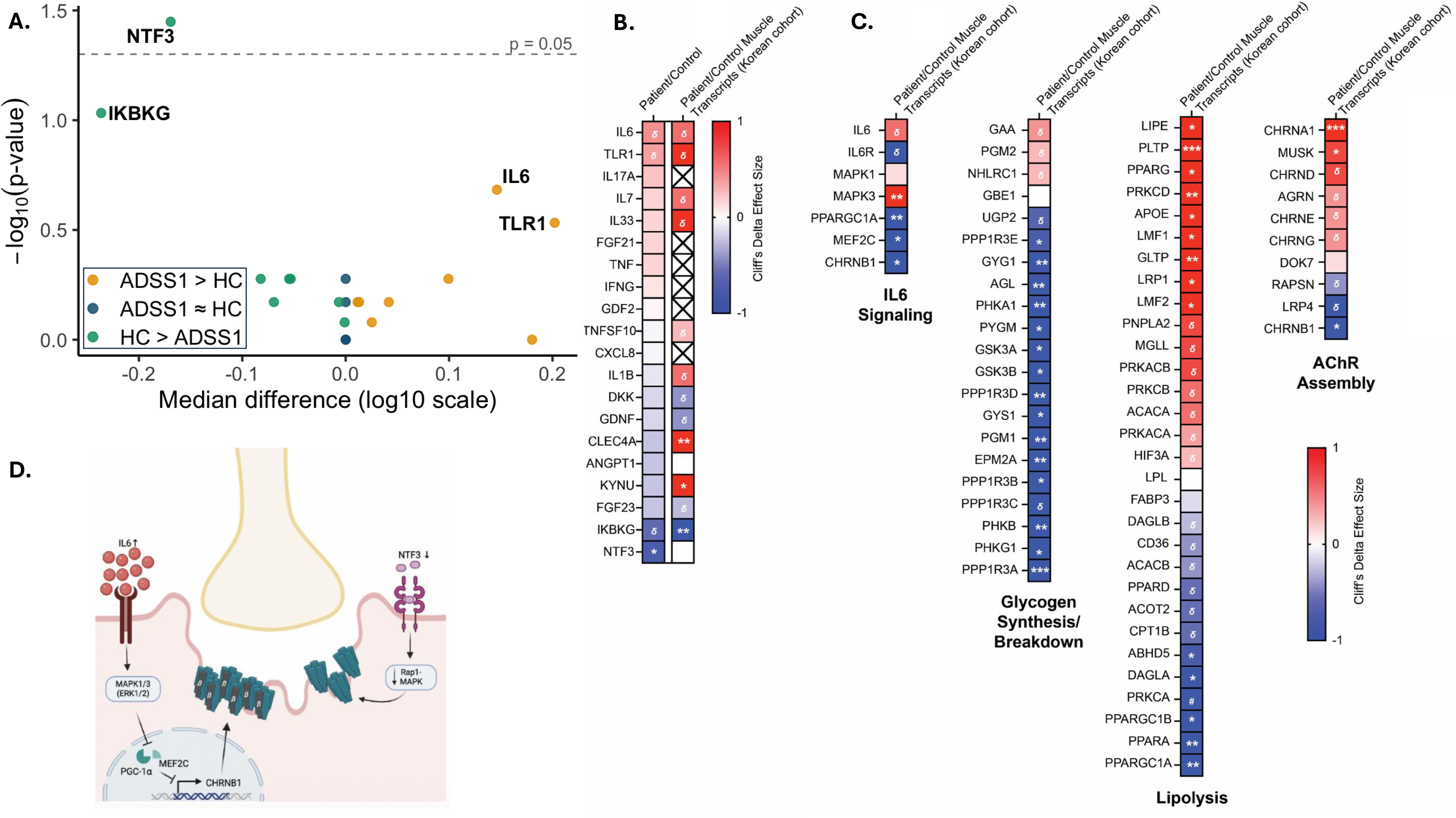
Plasma Proteomic Assessment. **(A)** Volcano plot depicting plasma proteomic group differences between patients with ADSS1 myopathy and HCs. The x-axis shows the observed difference in group medians for log10-transformed biomarker concentrations (ADSS1 − HC), while the y-axis shows −log10(raw p-values) from Wilcoxon rank-sum tests. Points are colored by direction of the observed group difference (ADSS1 > HC, HC > ADSS1, or approximately equal). The dashed horizontal line indicates p = 0.05. Effect magnitude and statistical significance are shown separately, as larger median differences do not necessarily correspond to smaller p-values in small exploratory samples. The uncorrected p-values reflect the consistency of rank ordering between groups, whereas median differences summarize central tendency. Markers with larger observed differences but greater within-group variability may therefore show weaker statistical evidence. See also **Supplemental Table 3. (B)** Muscle transcriptomic signature from Korean patients (n=4) and **(C)** IL-6 signaling, glycogen synthesis and breakdown, and acetylcholine receptor (AChR) assembly transcriptional pathways. An IL-6 and NTF3 driven neuromuscular junction modelling mechanism is proposed, where IL-6 activates MAPK1/3-ERK1/2 inhibition of transcription factors PGC1α (PPARGC1A) and MEF2C, and CHRNB1 transcription, which promotes ACHR clustering at the motor end plate. Reduced NTF3 signaling has a similar effect on AChR clustering via Rap1-MAPK signaling. Increase circulating IL-6 and reduced circulating NTF3 may be biochemical indicators of defective neuromuscular transmission in ADSS1 myopathy. Symbols denote: *p<0.05, **p<0.01, ***p<0.001, 8>0.3. **(D)** A proposed theoretical molecular model at the neuromuscular junction describing signaling pathways initiated by downregulated NTF3 and upregulated IL-6.

We subsequently probed an existing muscle transcriptomics data set derived from Korean ADSS1 myopathy patients to explore muscle-related expression of these proteins (**Figure 5B**).^3^ IL-6, TLR1 and ILBKG matched across data sets based on direction of regulation (IL-6 and TLR1 upregulated, IKBKG downregulated) and Cliff’s δ effect size > 0.2. Because IL-6 was consistently upregulated across data sets, we further probed the muscle transcriptome IL-6 signaling pathway (**Figure 5C**). Along with IL-6 transcriptional upregulation (p>0.05, δ=0.66), there was downregulation of soluble IL-6 receptor (IL-6R; p>0.06, δ=-1). There was evidence of downstream Mitogen-activated protein kinase 3 (MAPK3)-mediated inhibition of metabolic stress program transcription (Peroxisome proliferator-activated receptor gamma coactivator 1-alpha (PPARGC1A; p<0.01) and Myocyte-specific enhancer factor 2C (MEF2C; p<0.05) downregulation), including suppression of glycogen synthesis in favor of breakdown and glucose dominant metabolism. This mechanism has been linked to loss of neuromuscular junction (NMJ) stability and transmission via downregulation of CHRNB1-mediated acetycholine receptor (AChR) assembly. Muscle CHRNB1 was markedly downregulated in the Korean muscle transcriptome (p<0.05; **Figure 5C**). Reduced plasma NTF3 protein may indicate AChR clustering and NMJ loss since this protein is similarly necessary for AChR clustering (**Figure 5D**). Our data suggest muscle inflammation may exacerbate clinical phenotype, although it is unclear whether IL-6 emanates from muscle, intramuscular adipose tissue or immune cells, specifically.

### Associations Among Study Parameters

Spearman’s cross-correlation analysis combining data from ADSS1 myopathy and HC cohorts indicated that lower NTF3 levels associated with poorer performance on upper extremity motor tasks (9-hole peg test and grip strength) as well as higher upper extremity resistance and lower reactance and phase on EIM, (indicative of poorer muscle health) (**Figure 6A**). IL-6 and TLR1 showed opposite associative trends relative to NTF3 and IKBKG. Upper extremity resistance values associated with severity of self-reported mobility deficits. NTF3 and IKBKG also showed negative correlations with speech production parameters informing on vocal weakness (i.e., asthenia, breathy, or sustained vowel HNR and syllable repletion duration) **(Figure 6B)**.

**Figure 6:**
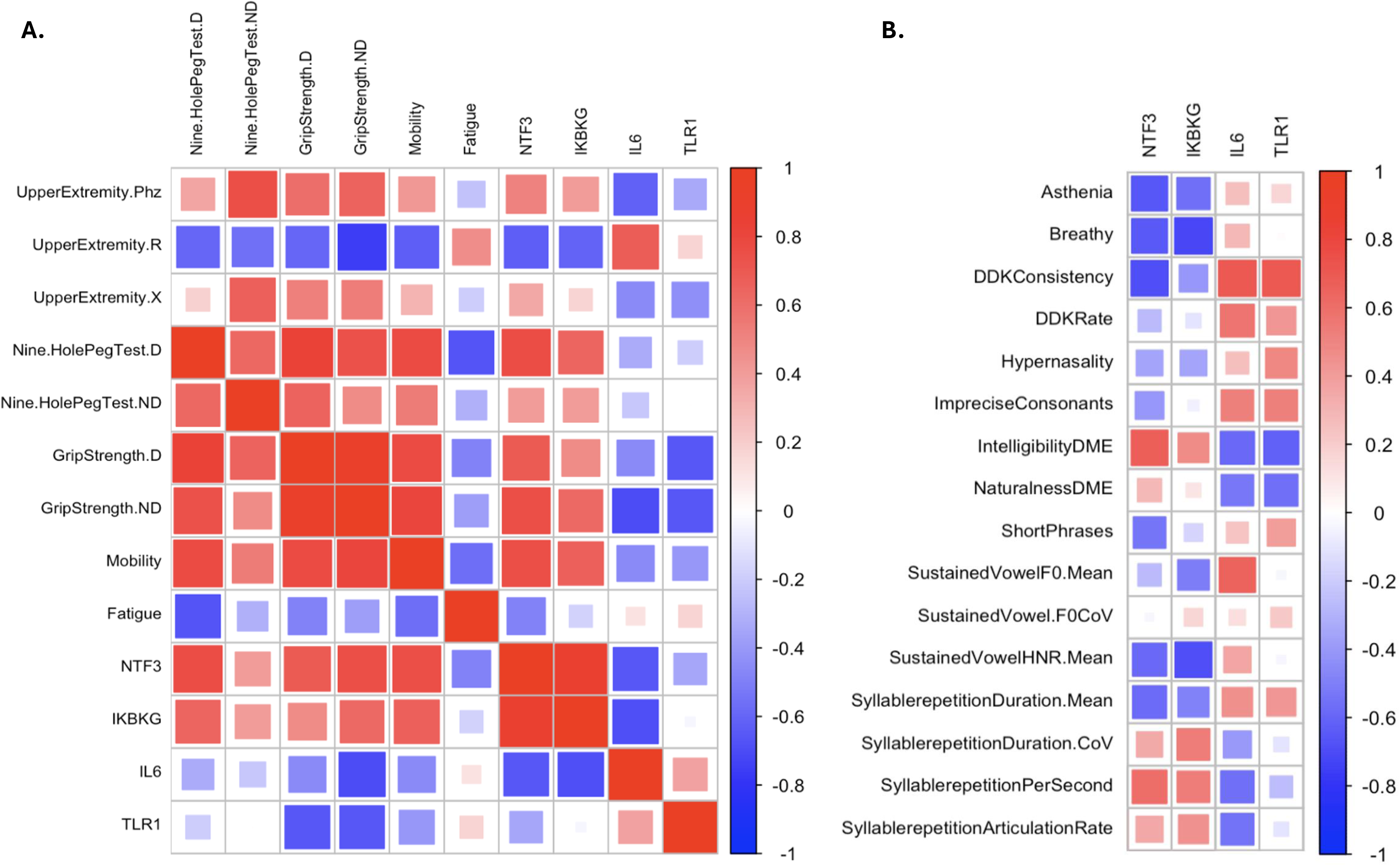
Biomarker Associations. Spearman’s cross-correlation analysis was performed integrating upper extremity EIM parameters (Phz: Phase; R: Resistance; and X: Reactance), performance on the 9-Hole Peg Test (D: Dominant; ND: Nondominant) and grip strength, self-reported measures of mobility and fatigue, and plasma proteomic markers **(A)**. In the latter, only proteins associated with a 8 (medium): 0.3–0.5 or 8 (large): > 0.5 effect sizes were included. A parallel Spearman’s cross-correlation analysis was performed incorporating plasma proteomic markers and perceptual and acoustic speech parameters **(B).**

## DISCUSSION

Patients with ultra-rare ADSS1 myopathy experience heterogenous disease trajectories, including age of onset, intensity of symptom progression and impact on quality of life, which complicates clinical care and prognosis^7^. In our recent multinational investigation of ADSS1 myopathy patients and caregiver experiences, decline in motor performance or mobility and muscle weakness were the most frequently reported symptoms impacting patients.^9^ Furthermore, ∼97% of patients reported fatigue which, impacts quality-of-life. While improving motor function and slowing disease progression were identified as critical treatment priorities for patients, limited and geographically disparate natural history data are a significant barrier to therapy development – especially the absence of validated biomarkers, functional endpoints and genotype-phenotype correlations. Here, we sought to quantitate potentially useful non-invasive biomarkers and functional endpoints in a small, singular timepoint deep phenotyping study of ADSS1 myopathy patients. Based on our previous pilot investigation of EIM in three patients with ADSS1 myopathy, we especially sought to validate EIM as a readily accessible, cost-effective and geographically portable tool for monitoring natural history in ADSS1 myopathy.^11^

The EIM findings obtained in the present study support the presence of a consistent and biologically meaningful electrical signature indicative of compromised muscle integrity in ADSS1 myopathy. Histopathological findings reported in muscle biopsy samples from patients with ADSS1 myopathy indicate marked disorganization of the muscle tissue microarchitecture, including abnormal alterations in muscle fiber architecture, increased interstitial and endomysial fibrosis, loss of intermyofibrillar organization, and adipose tissue accumulation in perimysial regions.^1,3,4,33^ Such microstructural abnormalities are biophysically plausible contributors to the abnormal electrical parameters observed in EIM measurements. Reductions in reactance are thought to reflect decreased tissue capacitance resulting from disruptions in cell membrane integrity and reductions in muscle fiber volume. Conversely, increases in resistance are likely associated with the heightened impedance to electrical current imposed by increased fibrotic and/or adipose tissue components within the muscle.^11^ The observation that EIM parameters change in a similar manner (i.e., increase in resistance and decrease in reactance and phase) in both upper and lower extremities suggests that muscle involvement in ADSS1 myopathy may represent a widespread and systemic process rather than a localized phenomenon. Comparable EIM patterns have previously been shown to reflect structurally and biophysically compromised muscle tissue across various neuromuscular disorders, and our findings demonstrate a high degree of concordance with the pilot study describing the application of EIM in ADSS1 myopathy. By extending these EIM observations and integrating them with multimodal assessments, our results further support the potential of EIM as a generalizable biomarker reflecting alterations in muscle composition in ADSS1 myopathy.^11,29,34,35^

In a prior study focusing on MRI-based analyses of lipomatous tissue in late-onset Pompe disease, muscle groups predominantly involving the posterior compartment were reported to be most severely affected.^36^ Although quantitative data regarding compartment-specific fat distribution in ADSS1 myopathy remain limited in the literature, our findings demonstrate that posterior compartment muscles also exhibit higher fat fractions in this cohort. Of note, all patients with ADSS1 myopathy who underwent musculoskeletal MRI were ambulatory. In the present study, MRI assessments were performed in only three patients, precluding statistically meaningful correlation analyses between MRI-derived fat fraction data and EIM measurements. Nevertheless, previous studies have demonstrated that changes in muscle composition—particularly increased fibrosis and fat infiltration—are associated with elevated resistance values in EIM. Accordingly, it is plausible that meaningful correlations between these modalities could be established in a larger ADSS1 patient population with more extensive MRI data.^29,35^ While MRI provides detailed, high-resolution structural information and remains a valuable modality for assessing muscle integrity, its routine clinical use is limited by high cost, substantial resource requirements, and operational constraints, particularly in individuals with restricted mobility.^37^ With further validation, EIM may prove to be a more practical alternative approach, as it is portable nature and relatively low cost allow for application across diverse clinical and non-clinical settings, including home- and community-based environments.^35^

Speech assessments further indicate that muscle involvement in ADSS1 myopathy is not limited to limb musculature but also extends to, for example, the oropharynx.^5^ Findings such as vocal asthenia, reduced speech intelligibility, and impaired voice quality are consistent with prior studies reporting prominent ADSS1 gene expression in oropharyngeal muscles.^5^ Within the context of neuromuscular disorders, speech characteristics have been shown to represent a functional manifestation of peripheral muscle weakness independent of central nervous system involvement and may therefore be affected even at early disease stages across various conditions. In this context, quantitative speech parameters appear to represent sensitive clinical indicators of functional disease burden in ADSS1 myopathy. Importantly, the feasibility of administering speech assessment batteries even in moderate to advanced disease stages provides a complementary functional window in situations where conventional motor function tests cannot be performed. When evaluated alongside EIM, speech analysis emerges as a component of a comprehensive, noninvasive approach for monitoring disease severity and functional impairment in ADSS1 myopathy. Furthermore, the absence of overt abnormalities on MRI of the masseter muscle in our patients raises the possibility that other muscle groups critical for speech—such as intrinsic tongue muscles, suprahyoid muscles, and pharyngeal constrictors, which are technically more challenging to assess using imaging or impedance-based methods—may be affected. However, direct and robust evidence supporting involvement of these muscle groups is currently lacking. Consequently, the application of speech assessments represents a particularly valuable strategy for capturing functional impairments that may be difficult to detect using structural evaluation methods alone.

The preservation of cognitive performance observed in this study supports the notion that ADSS1 myopathy primarily affects peripheral muscle and the neuromuscular junction. While previous studies based on patient and caregiver reports have described subjective cognitive complaints in some individuals—particularly involving attention, concentration, and memory—these symptoms have generally been reported as mild and not associated with substantial limitations in daily, academic, or occupational functioning. In this context, such cognitive complaints have been hypothesized to reflect secondary effects of the disease, including fatigue, motor limitations, and psychosocial burden, rather than primary central cognitive dysfunction.^9^ Consistent with these observations, our objectively derived cognitive testing results suggest that central cognitive functions are largely preserved in ADSS1 myopathy and that no disease-specific pattern of primary or widespread cognitive impairment is evident.

Although ADSS1 myopathy is caused by an inborn error of purine metabolism, overt metabolomic disruptions of muscle energy homeostasis are absent in ADSS1 mutant models, suggesting disease etiology is complex and multi-factorial.^38,39^ The canonical function of ADSS1 in AMP biosynthesis is well accepted in the literature.^40^ However, recent evidence implies ADSS1 has additional non-canonical roles by regulating epigenetic programmers (e.g., HDAC3) and NMJ assembly and transmission.^39,41^ These, as well as other undiscovered non-canonical mechanisms, may be protective to conserve cellular energy utilization at the expense of muscle function, and transmit unique protein signatures detectable in biofluids. Of the plasma proteins we assayed for biomarker potential, IL-6 was the most robust candidate being similarly upregulated in our plasma samples and muscle transcriptome.^3^ IL-6 is a highly prognostic biomarker in at least one blood disease and is a therapeutically amenable biomarker of inflammation in several others (e.g., rheumatoid arthritis, reviewed in).^42-44^ IL-6 is a pleiotropic cytokine, depending on the secreting tissue. As a myokine, it is a purported energy allocator released during muscle energy stress in response to metabolic signals, e.g., lactic acid accumulation and glycogen depletion, to mobilize energy stored in adipose tissue, skeletal muscle and the pancreas, and to stimulate liver gluconeogenesis. It simultaneously augments muscle uptake of mobilized energy by increasing insulin sensitivity and glucose transporter activity.^43^ In contrast, when secreted by immune cells in a specific cytokine suite (including IL-17, TNFα and IL-1β), IL-6 drives energy allocation to immune cells to support chronic inflammation. Plasma IL-6 concentration negatively associated with EIM phase (whole and upper body), non-dominant hand grip strength and plasma proteins, NTF3 and IKBKG, and positively associated with especially upper extremity EIM resistance in our study. Whether these signals reflect muscle adiposis driven pathology or are dependent on muscle IL-6-mediated signaling requires further investigation. IL-6 may be therapeutically amenable in ADSS1 myopathy based on preliminary evidence of anti-inflammatory efficacy in mutant zebrafish^45^.

Some limitations of this study should be acknowledged. First, due to the ultra-rare nature of ADSS1 myopathy, the sample size is inevitably limited, which constrains statistical power and underscores the need for validation of these findings in larger and more heterogeneous patient cohorts. Nevertheless, despite the small sample size, the consistency and magnitude of effects observed across EIM assessments suggest that the findings are meaningful. Second, the cross-sectional study design does not permit direct evaluation of disease progression or assessment of the longitudinal sensitivity of EIM, speech production parameters, or other study measures. Future longitudinal studies will be essential to clarify the capacity of EIM, MRI, motor testing, and speech measures to predict disease progression and monitor therapeutic responses in ADSS1 myopathy. Third, the 9-hole peg test could not be performed by one severely progressed individual, highlighting a limitation of function-based testing for assessing advanced disease. Muscle performance and range of motion were also not evaluated in the trunk or lower extremities nor in the context of functional mobility tasks (e.g., walk tests, motor performance batteries) and may provide valuable insight into disease progression and treatment efficacy in future studies involving ambulatory patients. Fourth, the limited number of patients undergoing MRI restricted comprehensive and statistically robust analyses of the relationship between EIM and muscle fat fraction. Moreover, although EIM provides a holistic reflection of changes in muscle composition, its ability to distinguish among specific pathological processes—such as fat infiltration, fibrosis, muscle atrophy, and loss of cellular organization—is inherently limited. As emphasized in other neuromuscular disorders, this underscores the importance of interpreting EIM findings in conjunction with imaging and functional measures. Finally, despite the substantial clinical value of speech assessments, the lack of direct imaging or EIM-based evaluation of the underlying muscle groups involved in speech (e.g., intrinsic tongue muscles, suprahyoid muscles, and pharyngeal constrictors) renders interpretations in this domain necessarily indirect. Nonetheless, this limitation does not diminish the potential of speech measures as sensitive functional outcome measures in ADSS1 myopathy; rather, it highlights the need for future multimodal approaches.

Despite these limitations, this study represents a comprehensive investigation integrating EIM, speech assessments, and functional measures in the context of the ultra-rare ADSS1 myopathy and provides a strong conceptual and methodological framework for future studies involving larger cohorts and longitudinal designs. The adoption of multimodal evaluation strategies is likely to play a critical role in identifying reliable biomarkers, enabling more sensitive tracking of disease progression, and facilitating the development of meaningful clinical endpoints for future interventional studies in ADSS1 myopathy.

## CONCLUSION

In conclusion, this study demonstrates that a multimodal, non-invasive assessment framework integrating electrical impedance myography, quantitative motor and speech evaluations, imaging, and plasma proteomics provides a sensitive and biologically informative approach for characterizing structural and functional disease burden in ADSS1 myopathy. Despite the inherent limitations of small sample size, the consistency of findings across complementary modalities supports the utility of this approach for refined phenotyping and objective monitoring in this ultra-rare disorder. These results lay an important foundation for future longitudinal and multicenter studies aimed at validating biomarkers, improving natural history characterization, and supporting the development of meaningful clinical endpoints for therapeutic trials in ADSS1 myopathy.

## AUTHOR CONTRIBUTIONS

All authors substantially contributed to the work and were involved in (a) conception and design of the study and/or analysis and interpretation of data, and (b) revising the article critically for important intellectual content. All authors approved the final manuscript as submitted and agreed to be accountable for all aspects of the work. MKY, RvG, HvdH and JU collected the data. MKY, RvG, NS, PYL, APV, WAH, and SBR designed the study. MKY, RvG, BSC, NS, GJ, CAT, JS, VR, EEH, CL, PYL, APV, ER, SBR, and JU analyzed data. MKY, RvG, GJ, APV, HJP, ER, and JU wrote the first version of the manuscript. MKY, RvG, WAH, SBR, and JU initiated this study. JU obtained funding for the study. All authors critically revised the manuscript for important intellectual content.

## ACKNOWLEDGMENTS

We are deeply thankful to the individuals living with ADSS1 myopathy and their families for their willingness to participate in this study and the Cure ADSSL1 for assistance with patient outreach.

## FUNDING INFORMATION

This study was funded by the Boston Children’s Hospital CHMC Anesthesia Foundation to JU.

## CONFLICT OF INTEREST STATEMENT

WAH and JU have received funding support from Sanofi Pharmaceuticals and Amicus Therapeutics. WAH is also on the advisory board for Sanofi Pharmaceuticals, Amicus Therapeutics, IntraBio Inc, Cyclo Therapeutics, Agyany Pharma, Azafaros, and Denali Therapeutics. SBR has equity in and serves a consultant and scientific advisor to Myolex, Inc., and Haystack Diagnostics, Inc., companies that design impedance devices for clinical and research use; he is also a member of the Myolex’s Board of Directors. The companies also have an option to license patented impedance technology of which SBR is named as an inventor. SBR is compensated for work on the company’s scientific advisory board and owns equity in the publicly traded company. APV is Chief Science Officer of Redenlab Ltd., which develops digital speech biomarkers used in this study. All other authors have nothing to disclose.

## DATA AVAILABILITY STATEMENT

The data that support the findings of this study are available from the corresponding author upon reasonable request.

## ETHICS STATEMENT AND INFORMED CONSENT

This study protocol was approved by the Boston Children’s Hospital Institutional Review Board (IRB-P00040784). All participants (and/or their legal guardians) gave written informed consent prior to participation.

## ANIMAL RIGHTS

This article does not contain any studies with animal subjects performed by any of the authors.

---

**Supplemental Figure 1.**
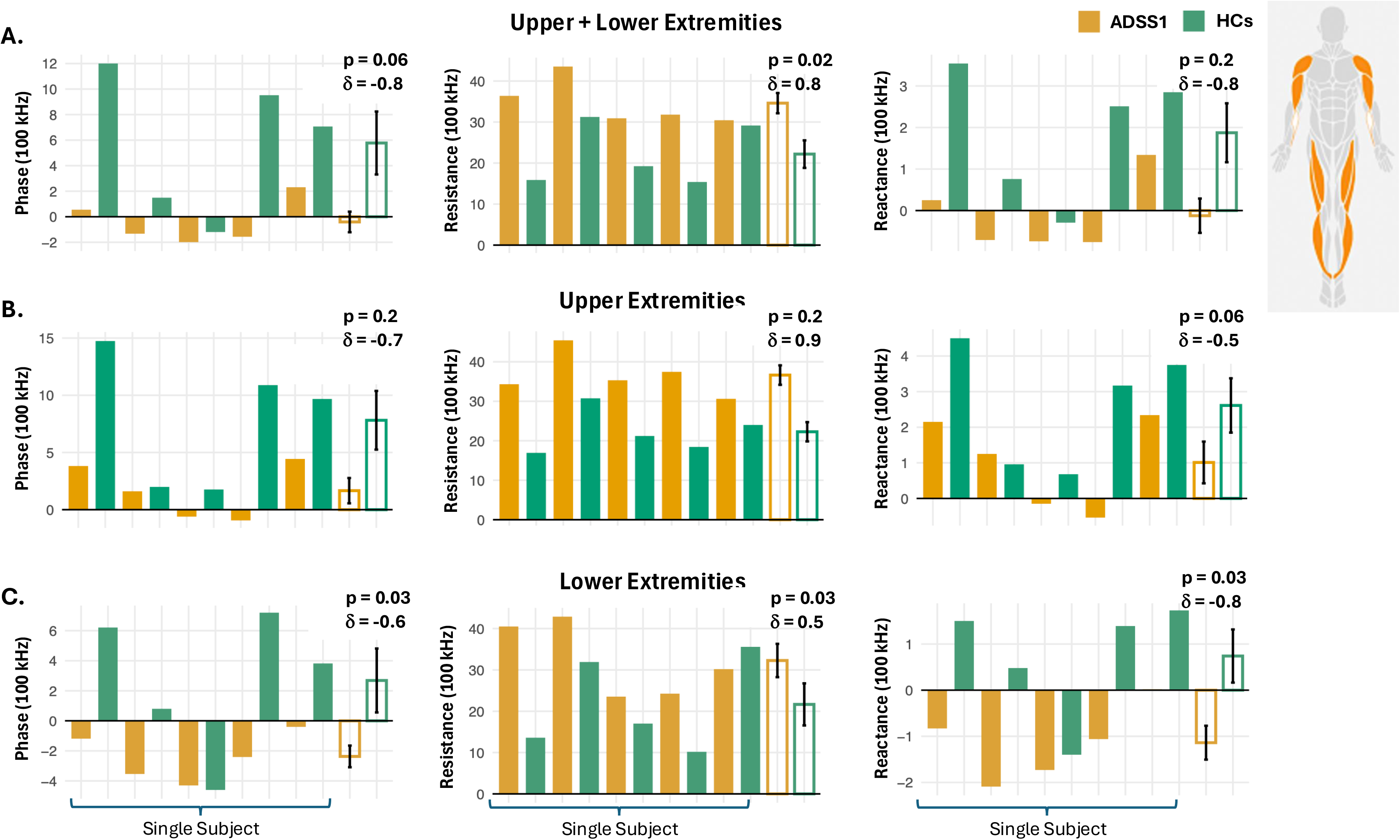

## Notes

### Author Declarations

This study protocol was approved by the Boston Children's Hospital Institutional Review Board (IRB-P00040784). All participants and or their legal guardians provided written informed consent prior to participation.

